# Systematic SARS-CoV-2 S Gene Sequencing in Wastewater Samples Enables Early Lineage Detection and Uncovers Rare Mutations in Portugal

**DOI:** 10.1101/2023.10.30.23297774

**Authors:** Ana C. Reis, Daniela Pinto, Sílvia Monteiro, Ricardo Santos, João Vieira Martins, Alexandra Sousa, Rute Páscoa, Rita Lourinho, Mónica V. Cunha

## Abstract

As the COVID-19 pandemic reached its peak, many countries implemented genomic surveillance systems to track the evolution and transmission of SARS-CoV-2. Transition from the pandemic to the endemic phase prioritized alternative testing strategies to maintain effective epidemic surveillance at the population level, with less intensive sequencing efforts. One such promising approach was Wastewater-Based Surveillance (WBS), which offers non-invasive, cost-effective means for analysing virus trends at the sewershed level. From 2020 onwards, wastewater has been recognized as an instrumental source of information for public health, with national and international authorities exploring options to implement national wastewater surveillance systems and increasingly relying on WBS as early warning of potential pathogen outbreaks. In Portugal, several pioneer projects joined the academia, water utilities and Public Administration around WBS.

To validate WBS as an effective genomic surveillance strategy, it is crucial to collect long term performance data. In this work, we present one year of systematic SARS-CoV-2 wastewater surveillance in Portugal, representing 35% of the mainland population. We employed two complementary methods for lineage determination - allelic discrimination by RT-PCR and S gene sequencing. This combination allowed us to monitor variant evolution in near-real-time and identify low-frequency mutations.

Over the course of this year-long study, spanning from May 2022 to April 2023, we successfully tracked the dominant Omicron sub-lineages, their progression and evolution, which aligned with concurrent clinical surveillance data. Our results underscore the effectiveness of WBS as a tracking system for virus variants, with the ability to unveil mutations undetected via massive sequencing of clinical samples from Portugal, demonstrating the ability of WBS to uncover new mutations and detect rare genetic variants.

Our findings emphasize that knowledge of the genetic diversity of SARS-CoV-2 at the population level can be extended far beyond via the combination of routine clinical genomic surveillance with wastewater sequencing and genotyping.

## INTRODUCTION

During the coronavirus disease 2019 (COVID-19) pandemic, detection of severe acute respiratory syndrome coronavirus 2 (SARS-CoV-2) in faeces accelerated the introduction of wastewater surveillance in a variety of settings, including cities, airports, hospitals, and university campuses (Agrawal et al., 2021; Medema et al., 2020; Nkambule et al., 2023). At the population level, wastewater-based surveillance (WBS) has been implemented solely as a complementary method to clinical surveillance (Peccia et al., 2020). However, WBS enabled correlating the real-time information of SARS-CoV-2 viral loads in sewage with the increase of positive cases and hospitalizations (Galani et al., 2022; Perez-Zabaleta et al., 2023), in addition to the identification of outbreak hotspots within a given region at the sewershed level (Fontenele et al., 2021; Medema et al., 2020; Monteiro et al., 2022). Moreover, WBS has also enabled detection of novel SARS-CoV-2 mutations in urban wastewater, which had not been detected by standard clinical surveillance (Karthikeyan et al., 2022; Smyth et al., 2022). WBS has thus become a public health instrument helping to track SARS-CoV-2 spread.

Given the potential contribution of both symptomatic and asymptomatic shedder individuals to SARS-CoV-2 viral load in sewage (Elsamadony et al., 2021; Foladori et al., 2020; Wu et al., 2020), wastewater surveillance presents a less biased and resource-effective approach for obtaining an overview of the viral diversity circulating in a community, in contrast to the currently dominant approach of clinical genomic surveillance. Therefore, in recognition of the immense potential of WBS, the European Commission recommended to Member States the implementation of regular SARS-CoV-2 monitoring through molecular analyses and sequencing of wastewater (*Commission Recommendation (EU) 2021/472*, 2021). Hence, a considerable number of studies with diverse aims, from establishing methodologies to finding new mutations, have reported results from WBS of European sub-populations (Agrawal et al., 2021; Amman et al., 2022; Galani et al., 2022; Izquierdo-Lara et al., 2021, 2023; Katharina et al., 2022; La Rosa et al., 2023; Medema et al., 2020; Monteiro et al., 2022; Pérez-Cataluña et al., 2022; Perez-Zabaleta et al., 2023). However, it is important to note that such studies are not limited to Europe; similar investigations have been conducted in American (Crits-Christoph et al., 2021; Fontenele et al., 2021; Gregory et al., 2022; Karthikeyan et al., 2022; Nemudryi et al., 2020; Peccia et al., 2020; Sapoval et al., 2023; Silva et al., 2022; Smyth et al., 2022; Swift et al., 2022) and Asian (Bar-Or et al., 2021) sub-populations as well. The data collected are of significant importance for comprehending methodological challenges and uncertainty sources, comparing the performance of WBS versus clinical surveillance, and extracting additional valuable information from WBS data.

The COVID-19 pandemic has been characterized by the sequential emergence of various variants of interest and variants of concern (VOC), with the most recent being Omicron (pangolin lineage B.1.1.529), initially identified in November 2021 (Rambaut et al., 2020; WHO, 2023). These VOCs are characterized by constellations of mutations that, to various degrees, appear to impact on virus transmissibility, clinical outcome, evasion from vaccine-derived adaptive immune responses, and effectiveness of diagnostic tests (Khateeb et al., 2021; Kuzmina et al., 2022; Tao et al., 2021). To date, wastewater analysis has routinely employed two approaches: (i) reverse transcription quantitative PCR (RT-qPCR) or reverse transcription-droplet digital PCR (RT-ddPCR) for SARS-CoV-2 detection and quantification; and (ii) high-throughput sequencing for monitorization of SARS-CoV-2 mutations and variants (Bar-Or et al., 2021; Crits-Christoph et al., 2021; Nemudryi et al., 2020). Although PCR-based wastewater surveillance enables the quick and highly sensitive detection and quantification of virus load in sewage, with the possibility to target specific viral mutations, it can provide limited information on which lineages and/or variants are circulating, because the number of diagnosing mutations has greatly decreased with the increase of viral genetic diversity concurrent with the expansion of the Omicron VOC. Tracking SARS-CoV-2 genetic diversity thus became essential to detect precociously the emergence of novel SARS-CoV-2 variants with potentially higher transmissibility and/or immune evasion properties, demanding rapid, cost-efficient, and accessible methods.

In this work, we aimed to monitor SARS-CoV-2 variant evolution in near-real-time and identify both major and low-frequency mutations. For that purpose, we present one year of systematic SARS-CoV-2 wastewater surveillance in Portugal, during the implementation of the European Commission (EC) Recommendation no. 2021/472, representing about 35% of the mainland population. We performed a combined approach, using a series of allele-specific RT-PCR assays targeting a panel of six Omicron associated mutations chosen to differentiate the most prevalent circulating lineages, and targeted Illumina sequencing of the Spike (S) gene of wastewater samples recovered in Portugal between May 2022 and April 2023.

## MATERIALS AND METHODS

### Study design, wastewater sampling and SARS-CoV-2 detection

The national surveillance system set up during the implementation of the EC Recommendation no. 2021/472 in Portugal aimed to cover a significant part of the Portuguese population. Given the dimension of the agglomerations on the national territory, the wastewater treatment plants (WWTPs) monitored for SARS-CoV-2 were selected based on facilities serving more than 100000 equivalent population and by excluding WWTPs with a strong industrial component. Monitorization focused on 14 WWTPs from the largest cities of the country, reflecting 34.2% of the mainland population. One litre of 24-hour composite raw wastewater samples was collected twice a week from 14 WWTPs located in the North (*number of WWTPs*, n=4), Center (*n*=2), Lisbon and Tagus Valley (LTV) (*n*=4), and Algarve (*n*=4) regions (Figure S1), with a population equivalent ranging from 113200 to 920000. Wastewater collection took place between May 2022 and April 2023 resulting in a total of 1339 samples.

Wastewater processing, RNA extraction, detection, and quantification were performed as previously described by Monteiro *et al*. 2022 (Monteiro et al., 2022). The SARS-CoV-2 viral load in each sample was estimated via viral RNA detection through RT-qPCR using the Charité assays: the E_Sarbecco assay targeting the envelope protein gene, the RdRp assay targeting the RNA-dependent RNA polymerase gene, and the N_Sarbecco assay targeting the nucleoprotein (Corman et al., 2020). For these, the one-step RT-qPCR assays AgPath-ID™ One-Step RT-PCR (Thermo Fisher Scientific, USA) were used as described before (Monteiro et al., 2022).

### Allelic discrimination analysis to detect Omicron associated mutations in wastewater samples

A set of six commercially available probe-based genotyping RT-PCR assays was used for the identification of mutations associated to specific Omicron sub-lineages. The panel of allele discrimination assays was continuously updated throughout the sampling period to accommodate variation within circulating Omicron sub-lineages, as assessed by clinical epidemiologic surveillance (Figure S2). These genotyping assays targeted specific amino acid changes, namely G339D, L452R, Q493R and T475K at S gene, D3N at M gene, and L11F at ORF7b (Table S1).

Genotyping assays were performed using the TaqMan SARS-CoV-2 Mutation Panel (Thermo Fisher Scientific, USA) following the manufacturer’s instructions. Each reaction consisted of 5 μL of wastewater RNA, mixed with 4X TaqPath™ 1-Step RT-qPCR Master Mix CG (Thermo Fisher Scientific, USA) and 40X TaqMan probes (Thermo Fisher Scientific, USA), in a final volume of 20 μL. Each assay included sequence-specific forward and reverse primers to amplify the target region and two TaqMan minor groove binder (MGB) probes with non-fluorescent quenchers (NFQ) to detect the PCR-amplification fragments: one FAM dye-labelled probe binds the mutated gene, and one VIC dye-labelled probe binds the wild-type SARS-CoV-2 gene.

PCR cycling was performed on a StepOne^TM^ Real-Time PCR System (Applied Biosystems^TM^), under the following conditions: reverse transcription at 45°C for 15 minutes, Taq polymerase activation at 95°C for 2 minutes, followed by 45 cycles of denaturation at 95°C for 15 seconds, and annealing at 58°C for 45 seconds, with a final post-read for 30 seconds at 60°C. All RT-PCR experiments included a no-template control (NTC) (*i.e.*, DNase/RNase free water), and a wild-type AcroMetrix Coronavirus 2019 (COVID-19) RNA control (RUO) (Thermo Fisher Scientific).

Allelic discrimination results were then visualized on a scatter plot, contrasting reporter dye fluorescence (*i.e.*, mutated allele versus wild type allele), and analyzed using QuantStudio Design and Analysis Software version 2.5, via the genotyping analysis module with real-time data.

### S gene sequencing and reference-based analyses

From the complete set of samples, 332 were selected for S gene sequencing. These corresponded to two samples per month and per WWTP, as defined in the EC Recommendation. Whenever possible, the samples selected for sequencing were those that exhibited RT-qPCR amplification of at least two SARS-CoV-2 genes (E, N, or RdRp), with threshold cycle (Ct) values below 36, which were found to yield more genome coverage. Samples with higher Ct values were preferentially avoided to ensure best quality sequencing results and facilitate accurate SARS-CoV-2 variant/lineage assignment.

The RNA from the wastewater samples was purified with RNeasy MinElute Cleanup kit (Qiagen), following the manufacturer’s instructions, to eliminate impurities and inhibitors that could potentially interfere with the subsequent sequencing process. Total RNA was then quantified using the Qubit RNA HS Assay kit (Thermo Fisher Scientific, USA), following the manufacturer’s instructions. Samples with RNA concentrations above 20 ng/μl were selected for SARS-CoV-2 sequencing.

The SARS-CoV-2 target amplicon libraries were constructed using ARTIC SARS-CoV-2 RNA library (Eurofins, Konstanz, Germany), which was especially designed to amplify the S gene. Sequencing occurred using the Illumina NovaSeq 6000, according to the manufacturer’s specifications, with the paired-end module (2×150 bp) attachment. Eurofins sequencing strategy was optimized for the S gene sequencing in wastewater samples, with the application of 49 primers, including 22 primer pairs with 5 alternative primers to account for variability in the primer binding region. The 22 amplicons ranged in size from 200 to 250 base pairs, and collectively cover the entire S-gene sequence, spanning 3822 base pairs.

Read processing, reference-based alignment and variant analysis were performed using bioinformatics tools available in the Galaxy EU server (Afgan et al., 2018). Quality control reports were generated with FastQC (Galaxy version 0.11.9) using default parameters. Raw reads were cleaned up from adaptors (-ILLUMINACLIP) and low quality nucleotides (-SLIDINGWINDOW:4:30) using Trimmomatic (Galaxy version 0.38) (Bolger et al., 2014). Quality trimmed reads were aligned to the SARS-CoV-2 reference genome (NCBI accession number NC_045512.2) using the Burrows-Wheeler Aligner (Galaxy version 0.7.17) (Li, 2013) with default parameters; and reads with less than 35 bp were removed, using BAM filter (Galaxy version 0.5.9) in order to eliminate primers and short uninformative reads. Variant calling was performed using iVar pipeline (Galaxy version 1.4.2) (Grubaugh et al., 2019), with the minimum quality score threshold to count a base being set to 20, and the minimum frequency threshold to 0.03. The minimum number of reads to consider major variants (polymorphism with a frequency above 0.5) was set to 10, while for minor variants (polymorphism with a frequency between 0.03 and 0.49) the number of supporting reads was set to 20.

The nucleotide polymorphisms were evaluated considering their effect on their protein product by SnpEff (Galaxy version 4.5covid) (Cingolani et al., 2012). Finally, the quality metrics of the assemblies were obtained with Qualimap BAMQC (Galaxy version 2.2.2) (Okonechnikov et al., 2016).

Information concerning the worldwide occurrence of specific SARS-CoV-2 genetic polymorphisms and frequency was obtained from outbreak.info (Gangavarapu et al., 2023).

### SARS-CoV-2 lineage assignment

To perform SARS-CoV-2 lineage assignment, Freyja pipeline (Karthikeyan et al., 2022), currently available at https://github.com/andersen-lab/Freyja, was used. Freyja is a bioinformatics pipeline designed to estimate the relative abundance of SARS-CoV-2 lineages in a mixed sample. To assign a lineage to each sample, Freyja stores the single nucleotide polymorphism (SNP) frequencies for each of the lineage-defining mutations and recovers relative lineage abundance by solving a depth-weighted least absolute deviation regression. Note that greater sequencing depth estimate mutation frequencies more accurately, which results in more precise lineage assignments. Freyja assigns SARS-CoV-2 lineages based on the outbreak.info curated lineage metadata file that summarizes lineages by World Health Organization (WHO) designation.

### Data analysis

SARS-CoV-2 lineage assignment and mutation data derived from clinical surveillance were obtained from GISAID (https://www.gisaid.org/, accessed on the 20^th^ of June 2023) (Elbe and Buckland-Merrett, 2017; Shu and McCauley, 2017). Clinical samples from Portugal, with collection dates ranging between the 19^th^ of April 2022 and the 12^th^ of May 2023, comprehending the wastewater sample collection period plus two weeks before and afterward, were selected. This dataset was further filtered to samples isolated in the Health Regions covered by the WWTPs under monitoring in this study (*i.e.*, North, Center, Lisbon and Tagus Valley, and the Algarve) (Figure S1).

Data analysis and visualization was performed with custom Python 3.9.10 (Van Rossum and Drake, 2009) and R version 4.1.1 (R Core Team, 2021) scripts. Phyton modules pandas version 1.4.1 (McKinney, 2010) isoweek version 1.3.3, outbreak_data version 1.0.1, and numpy version 1.22.3 (Harris et al., 2020) were used for data manipulation. R packages dplyr version 1.1.2 (Wickham et al., 2023a), ggplot2 version 3.4.2 (Wickham, 2016), gridExtra version 2.3 (Auguie and Antonov, 2017) gtable version 0.3.3 (Wickham et al., 2023c), readr version 2.1.4 (Wickham et al., 2023b) BiocManager version 1.30.20 (Morgan and Ramos, 2023) drawProteins version 1.14.0 (Brennan, 2018), ggforce version 0.4.1 (Pedersen, 2022) and cowplot version 1.1.1 (Wilke, 2020) were used to create the visualizations in RStudio version 1.4.1717 (RStudio Team, 2021). Spike protein annotations were obtained from UniProt (accession number: P0DTC2; (The UniProt Consortium, 2023)); mutation occurrence per country was retrieved from outbreak.info (Gangavarapu et al., 2023) using the R package outbreakinfo version 0.2.0 (Alkuzweny et al., 2023); the number of sequences made available per country was retrieved from GISAID (Elbe and Buckland-Merrett, 2017; Shu and McCauley, 2017); and the Simpson evenness index was calculated using R package abdiv version 0.2.0 (Bittinger, 2020).

## RESULTS

### The adaptive combination of RT-PCR allelic discrimination assays successfully captures the co-occurrence and progression of the main Omicron sub-lineages

All the 1339 wastewater samples under focus in this study were tested for the presence of specific mutations associated to one or several Omicron sub-lineages. Given the fast evolution of Omicron sub-lineages, the panel of applied genotyping assays was regularly updated (Figure S2) throughout the sampling period to discriminate new sub-lineages. This meant adding new assays targeting diagnostic mutations of new sub-lineages and halting assays that no longer had discrimination power over the sub-lineages circulating in that period. These decisions were informed both by the results that were being obtained weekly, as well as by the clinical surveillance data made publicly available by the Directorate-General of Health (DGS) and the National Institute of Health Doutor Ricardo Jorge (INSA, 2021).

At sampling starting point, four genotyping assays were applied. The genotyping assay that detected a signature mutation of the Omicron VOC (*i.e.*, S:G339D) was selected to assess the circulation of older VOCs that possess the wild-type allele. Note that at the beginning of the sampling period, Omicron was already the dominant VOC. With the exception of ISO week 24, the mutant allele was exclusively detected until September 2022 (ISO week 38), from which point both alleles were regularly identified (Figure S3). Clinical surveillance data for all analyzed regions (Figure 1A) showed that, at that time point, the resurgence of older VOCs was not being observed, but instead detection of the wild type allele corresponded to the emergence of Omicron sub-lineages without this mutation, among them the recombinant sub-lineages of CH and XBB (named hereafter CH.X and XBB.X). Given that this mutation was no longer a signature mutation for the Omicron VOC, we suspended the application of this assay in the beginning of November 2022 (week 45).

**Figure 1:**
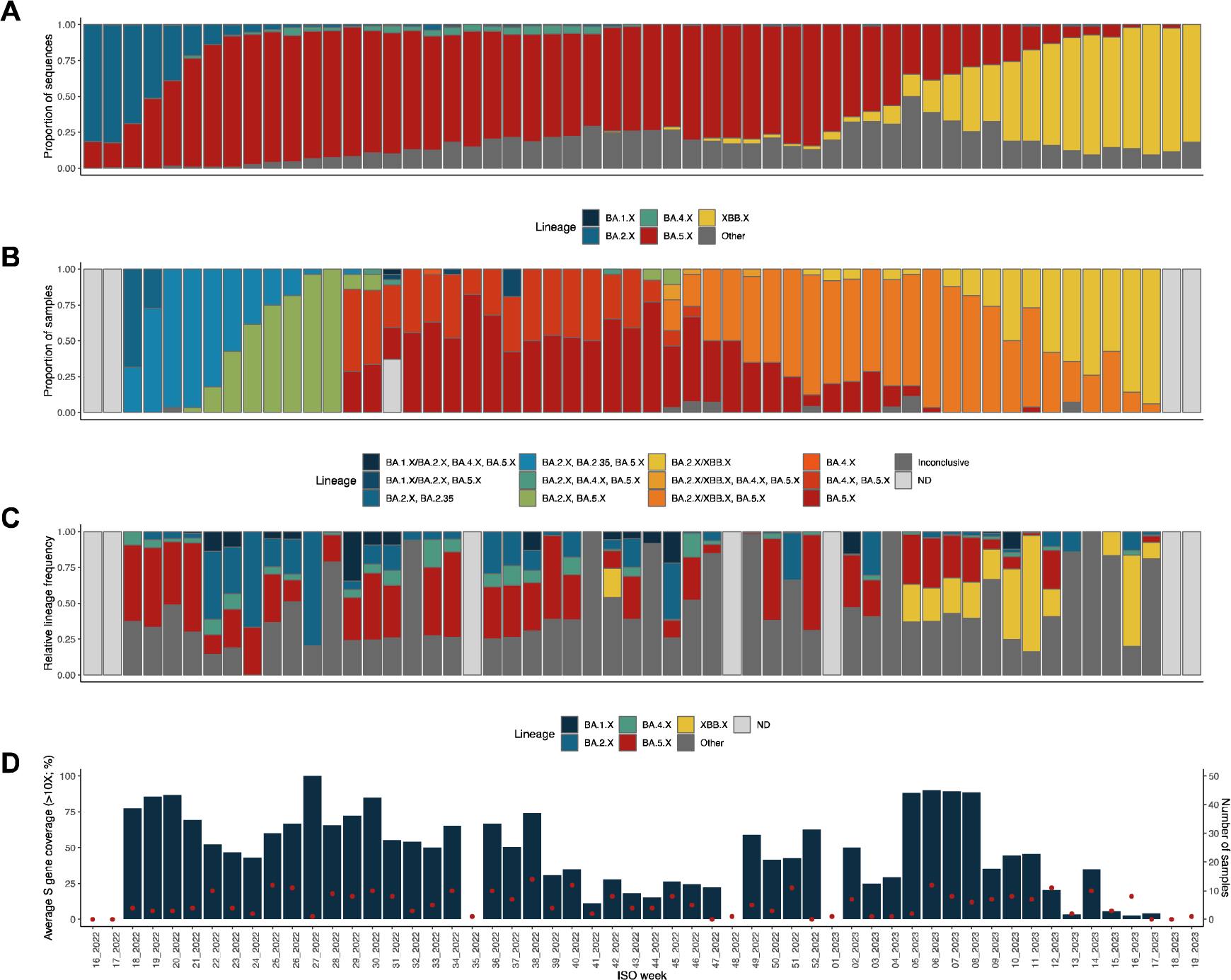
Lineage frequency evolution. **A.** Lineage assignment of sequences derived from clinical genomic surveillance. The samples were collected in Portugal (in North, Center, Lisbon and Tagus Valley, and Algarve Health Regions) between ISO week 16 of year 2022 and ISO week 19 of year 2023. Lineages were grouped with all descendant lineages of the five major circulating sub-lineages (BA.1, BA.2, BA.4, BA.5 and XBB) grouped into a single category. The remaining (minor) lineages were grouped into a single category (Other). **B.** Lineage assignments in WWTP samples based on real-time PCR diagnostic data (allelic discrimination assays). Based on the tested alleles, the samples were grouped into 14 categories. Most of the categories group samples in which multiple major circulating lineages were detected. The forward slash indicates that the test cannot distinguish between the two lineages. In a small number of samples, amplification was not obtained in all necessary real-time PCR assays and hence they were grouped in the category “Inconclusive”. Samples not tested were grouped in the Not Determined (ND) category. **C.** Lineage assignments in WWTP samples based on sequencing data. The data was grouped in the same categories as those in panel A for convenience of comparison. **D.** Average horizontal S gene sequencing coverage and number of sequenced samples per week (referring to the data presented in panel C). Vertical blue bars represent the average horizontal coverage of that week’s samples, and red dots represent the number of sequenced samples on that week. Note that ISO weeks 16 and 17 of 2022 and ISO weeks 18 and 19 of 2023 were out of our wastewater sampling period. On ISO week 35 of 2022, there was no sampling at all, and on ISO weeks 48 of 2022 and 01 of 2023, no samples were sequenced due to the low quality of the samples’ RNA.

Another assay applied at the beginning of our sampling period was able to identify the signature mutation of the Omicron BA.1 lineage and its daughter lineages (BA.1.X; S:T475K; Figure S2). However, we have only applied this assay during the first two weeks of the sampling period (weeks 18 and 19) given that the mutant allele was never amplified in any sample (Figure S3). This observation supported the hypothesis that BA.1.X sub-lineages were no longer circulating in the analyzed communities, which agreed with the clinical epidemiological data (Figure 1A) and led to the early exclusion of this assay from the panel. Between May 2022 (week 18) and November 2022 (week 48) the genotyping assay that identifies the S:Q493R mutation present in BA.1.X, BA.2.X and BA.3.X Omicron sub-lineages was applied (Figure S2). Knowing that BA.3.X had only been detected twice in Portugal through genome sequencing and before the start of our sampling period, and also being aware that BA.1.X was being reported from week 16 in percentages bellow 1%, the identification of the mutant allele S:Q493R was presumed indicative of BA.2.X circulation. Between ISO weeks 18 and 29, the clinical data shows the substitution of BA.2.X by BA.5.X (Figure 1A). BA.2.X sub-lineages were dominant in the clinical data until week 19, representing over 50% of the available sequences. In wastewater samples, the mutant allele of S:Q493R was detected in over 50% of samples for five weeks longer (*i.e.*, until week 24) but also with a decreasing tendency (Figure S3). Interpretation of results for samples with amplification of the mutant allele only, without amplification of the wild type allele, was translated as the presumable presence of 100% BA.2.X sub-lineages in the wastewater samples. Detection of this mutant allele stopped at week 20, after which amplification of both alleles was obtained, suggesting the coexistence of both BA.2.X and BA.5.X sub-lineages in the same sample. Similar results were observed at regional level, with the mutant allele of S:Q493R amplified in over 50% of samples until week 23 in Algarve, and until week 24 in North, Center and, Lisbon and Tagus Valley (LTV) regions (Figure S4). The identification of samples only with the amplification of the mutant allele stopped at week 19 in the North, Center and LTV regions, and at week 20 at Algarve. Despite the similarities, the increase in detection of the wild type allele, and consequently the decrease of BA.2.X detection, was faster in the LTV region, followed by Algarve, North and Center regions (Figure S4).

The mutation S:L452R is present in BA.4.X, BA.5.X and BA.2.35, and was also applied from the beginning of our sampling period and until week 29. Since the percentage of BA.4.X sequences isolated until week 29 was residual, the presence of the mutant allele was taken as an indication of the presence of BA.5.X or BA.2.35 sub-lineages. The combination of results from genotyping assays S:Q493R and S:L452R resolved the ambiguity of the assay for mutation S:Q493R in lineage assignment and allowed detecting the presence of BA.2.35 sub-lineage. Based on this, sub-lineage BA.2.35 was presumably present in Portugal at least until week 24.

At week 29, two new assays were added (Figure S2): the genotyping assay ORF7b:L11F, which identifies a signature mutation of BA.4.X sub-lineage, that was applied until the end of 2022 (week 52); and the genotyping assay M:D3N, which identifies the signature mutation of BA.5.X, that was applied until Abril 2023 (week 17).

Concerning BA.4.X sub-lineage, none of the analyzed samples revealed the unique amplification of the mutant allele, indicating that the presence of BA.4.X was concomitant at all times with BA.5.X. From week 29 until week 34, the signature mutation of BA.4.X was identified in most samples. At the national scale, the last reported detection of this lineage on clinically derived sequences is dated from week 48, while in wastewater samples the mutant allele was amplified until week 49. Considering the regional stratification, the identification of BA.4.X in wastewater continued to be reported later in time when comparing with clinical data (Figures 1, S4 and S5), except for the Center region. Specifically, at the Algarve, BA.4.X was last identified in wastewater at week 46, while in clinical data was at week 40; at LTV it was last reported in wastewater at week 49, while at clinical data was at week 43; at North, it was detected up to week 45 in wastewater and at week 43 in clinical data (Figure S5). The center region clinical data reported BA.4.X until week 48, while in wastewater it was detected up until week 46. Since the mutant allele was not detected at wastewater samples in the last three weeks of 2022, we suspended the implementation of the genotyping assay ORF7b:L11F in the end of 2022.

Regarding BA.5.X, the mutant allele (*i.e.*, M:D3N) was identified in all samples recovered between week 29 of 2022 and week 17 of 2023 (Figure S3), attesting the uninterrupted circulation of BA.5.X. This is in agreement with the clinical data that shows BA.5.X with an increased and dominant tendency until the end of 2022, when XBB.X started increasing their frequency (Figure 1). At the regional level, the scenario is similar, with the mutant allele amplified in over 50% of samples until week 13 of 2023 at North region, until week 12 at national level and at LTV region, until week 10 at Center region, and until week 9 at Algarve region (Figure S4). This shows that the substitution of BA.5.X by XBB.X was faster at Algarve region, followed by Center, then LTV and later in the North region.

Figure 1B presents a comprehensive summary of the wastewater data, enabling the easy comparison between the results from RT-PCR genotyping assays and the clinical data presented in Figure 1A. The samples are grouped by week, and the categories represent all the sub-lineages found in each sample. Note that, for 21 samples, the allele discrimination analysis was inconclusive due to lack of amplification, which might be due to the complex nature of wastewater samples that can contain PCR inhibitors and other contaminants. Another possible reason for the lack of amplification in these samples concerns the sensitivity of the assays. In fact, the manufacturer recommends the assays to be applied to samples whose Ct was inferior to 30 in the RT-qPCR for SARS-CoV-2 identification, which was not the case for any of these samples.

### S gene sequencing exhibits high sensitivity to sample quality variations

From the initial pool of 1339 samples, 332 were selected for S gene sequencing based on specific criteria: RNA concentration above 20 ng/µl, Ct values of the RT-qPCR for SARS-CoV-2 detection below 36, and the inclusion of two samples per month per WWTP. Given the pivotal role of the viral Spike protein in the infectious process, mediating viral entry into host cells (Jackson et al., 2022) and its significance in generating new VOCs with heightened transmissibility (Markov et al., 2023), targeted sequencing of the S gene was carried out.

The resulting data underwent meticulous analysis to identify diagnostic mutations, low-frequency mutations, and even new mutations, along with lineage assignment, facilitating direct comparison with data from clinical surveillance and the data generated in parallel via the RT-PCR genotyping assays.

Throughout the analyses, it became evident that the horizontal coverage of the S gene varied considerably, significantly impacting the number of mutations detected per sample (compare Figure 2A with Figure 2C). On average, the S gene’s horizontal coverage by at least 10 reads was 50.2 ± 29.5%, with 31% of the samples (n = 103) showing coverages above 70%. However, during specific time points, particularly in weeks encompassing October and November 2022, and January, March, and April 2023, the horizontal coverage dropped to an average of less than 25% of the S gene (Figure 2C). Lower coverages also coincided with higher values of the Simpson Evenness Index. A Simpson Evenness Index close to 1 indicates that all mutations have equal abundances in the set of samples of that week, while a value close to 0 suggests the sample set is dominated by only a few highly prevalent mutations. In our dataset, we observed that lower horizontal coverage of the S gene led to the detection of fewer mutations per sample, all with more comparable abundances in the sample set. On the contrary, higher horizontal coverage of the S gene resulted in the detection of a greater number of mutations per sample, with a few mutations being over-represented in the samples set. This observation suggests that, in some samples with low horizontal coverage of the S gene, we might not have been able to capture the full scope of mutations present.

**Figure 2:**
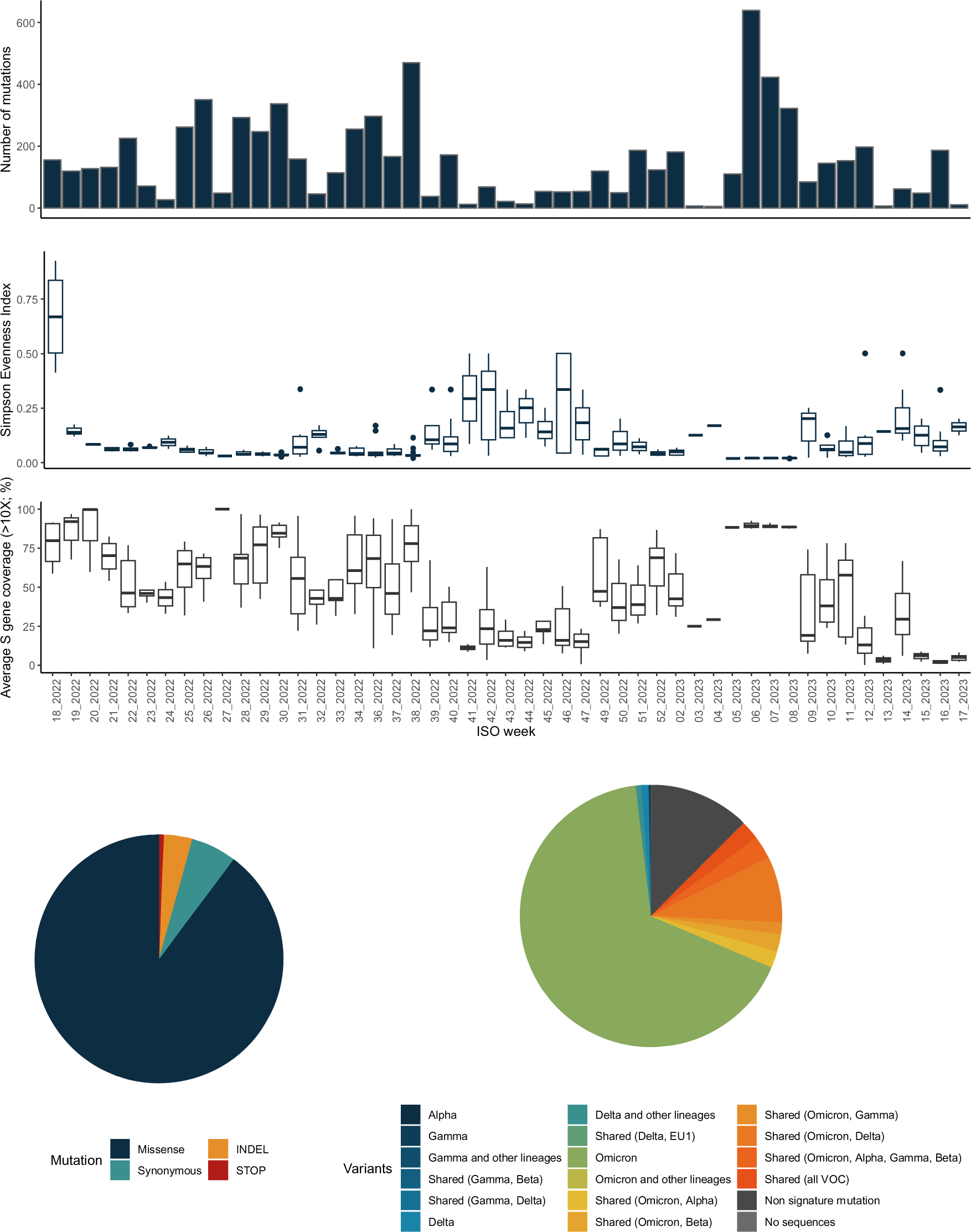
Number and type of S gene mutations. **A.** Number of S gene mutations by week. **B.** Box plot of the distribution of the Simpson Evenness Index calculated for each analyzed sample and represented by week. **C.** Distribution of the average horizontal S gene sequencing coverage determined for each sample and represented by week. Horizontal coverage of S gene of at least 10X is represented in percentage. **D.** Pie chart showing the relative proportion of each type of mutation detected in the samples. STOP refers to a mutation that generates a premature stop codon. **E.** Each detected mutation was assigned as being associated with one or multiple lineages based on occurrence. Two novel unique mutations were not assigned to any lineage (“No sequences”).

We then attempted to correlate the observed low horizontal coverage of the S gene, particularly evident in certain weeks (*e.g.*, weeks 39 to 47; Figure 2C), with various wastewater sample parameters such as the pH, biological oxygen demand, daily temperature, daily rainfall, and nitrogen and phosphorus concentrations. However, no significant correlations were found that could explain the observed low S gene coverage (data not shown). Similarly, no correlations were observed with RNA concentration or viral load, indicating that other factors might be at play, such as RNA integrity or the presence of PCR inhibitors commonly found in wastewater samples (Amman et al., 2022; Baaijens et al., 2021; Silva et al., 2022), despite our additional RNA purification steps. These factors can adversely affect the sequencing process, leading to limited sequence coverage and subsequently restricting downstream analysis.

### The majority of detected mutations are missense SNPs associated with the Omicron VOC

In the 332 sequenced samples, a total of 7168 SNPs and 278 insertions/deletions (INDELs) were discriminated within the S gene over time when compared to the SARS-CoV-2 reference genome (NC_045512.2). On average, each sample registered 22 ± 16 polymorphisms (comprising both SNPs and INDELs). Among these, the majority were missense SNPs (89.8%), followed by synonymous SNPs (5.9%), INDELs (3.7%), and the introduction of premature STOP codons (0.6%) (Figure 2D).

The identified missense SNPs were classified based on their occurrence within various VOCs. Some missense SNPs were associated with one or multiple VOCs, while others were not specifically linked to any known VOC or had never been previously reported (Figure 2E). As expected, most missense SNPs were associated with the Omicron VOC (Figure 2E), which was the dominant VOC in our sampling period. In addition, mutations associated with older VOCs such as Alpha, Delta, Beta, and Gamma were also sporadically observed in our samples (Figure 2E). Clinical-derived reports have Delta accounting for 0.01% of worldwide recovered SARS-CoV-2 sequences during our sampling period (source: GISAID on the 1^st^ of August 2023). Mutations associated with other VOCs, excluding Delta and Omicron, were sporadic, likely due to their very low prevalence, collectively representing only 0.002% of worldwide recovered sequences in our sampling period. Note that in the clinical surveillance data, other VOCs were never detected, which is not surprising given their residual circulation.

### Frequency and coverage constrain SNP detection in wastewater samples

We proceeded to compare the missense SNPs identified in our wastewater samples with those detected in clinical samples obtained during the same time period. The aim was to assess the performance of our wastewater analyses compared to clinical genomic surveillance. In this analysis, we focused on missense SNPs and found that only 10.7% of them were identified in both sample types - clinical and wastewater (Figure 3A). Most missense SNPs (61.8%) were exclusively found in clinical-derived sequences, while 27.4% were exclusively detected in wastewater samples (Figure 3A).

**Figure 3:**
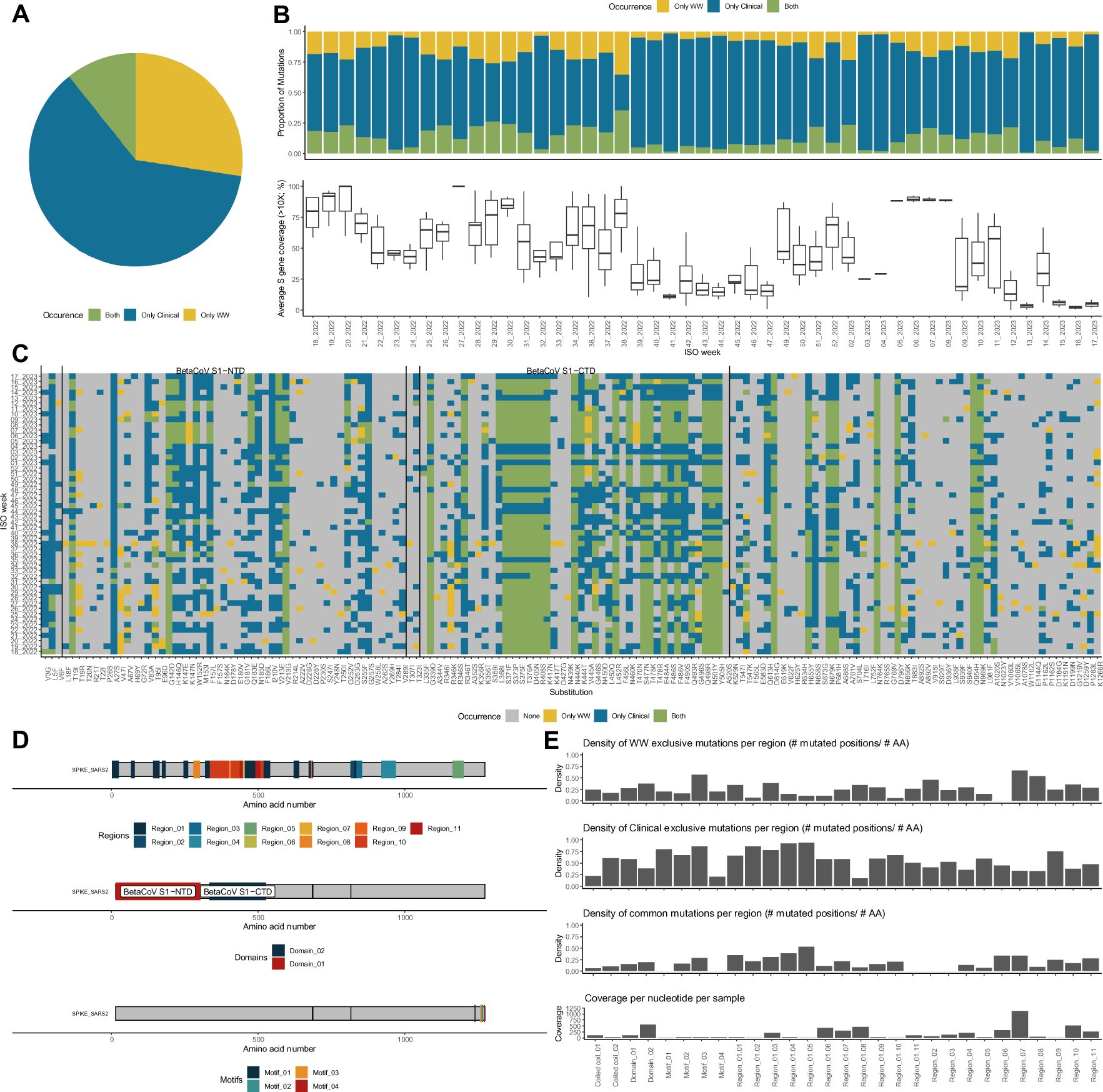
Occurrence of specific missense substitutions. A. The pie chart shows the relative proportions of substitutions found only in wastewater (WW) samples, only in Clinical samples and in both. B. The top bar plot shows how the relative proportion of missense substitutions found only in WW samples, only in Clinical samples or in both varies per week. The bottom box plots show the distribution of S gene horizontal coverage as in Figure 2C. C. For the set of substitutions that were found in both sample types (i.e., Clinical and WW), the occurrence per week in one of the samples, both or none is shown. D. Spike protein annotation with the Regions, Domains and Motifs analyzed in panel E. E. For each Spike protein Region, Domain, Motif and Coiled coil, the average coverage and the density of mutated positions only found mutated in WW, Clinical, or in both samples, are shown. Region_01, multiple occurrences of disordered region. Region_02, putative superantigen; may bind T-cell receptor alpha/TRAC. Region_03, receptor-binding domain (RBD). Region_04, integrin-binding motif. Region_05, receptor-binding motif; binding to human ACE2. Region_06, immunodominant HLA epitope recognized by the CD8+; called NF9 peptide. Region_07, putative superantigen; may bind T-cell receptor beta/TRBC1. Region_08, fusion peptide 1. Region_09, fusion peptide 2. Region_10, heptad repeat 1. Region_11, heptad repeat 2. Domain_01, BetaCoV S1-NTD. Domain_02, BetaCoV S1-CTD. Motif_01, binding to host endocytosis trafficking protein SNX27. Motif_02, diacidic ER export motif (host COPII). Motif_03, binding to host plasma membrane localizing/FERM domain proteins. Motif_04, KxHxx, ER retrieval signal (COPI).

Upon closer examination of this distribution, we observed that in weeks where we obtained higher horizontal coverage of the S gene, the percentage of mutations exclusively found in clinical samples decreased (Figure 3B).

We went deeper into our analysis by examining the consistency of detection of missense mutations present both in clinical and wastewater samples, aiming to identify any patterns throughout the sampling period and across the full extent of the Spike protein (Figure 3C). We observed three distinct sets of mutations. Firstly, there were mutations that occurred sporadically in both sample types, such as D228G, K417T, and S929T. These mutations are of low prevalence, both in Portugal and worldwide (below 1% prevalence as of August 1^st^, 2023, source: outbreak.info). Therefore, their detection in wastewater samples is also sporadic and not simultaneous with clinical detection. Secondly, there were mutations, such as S371F, N501Y, and Q954H, that were consistently found in both sample types throughout the sampling period. These mutations are high frequency mutations (above 50% prevalence worldwide and in Portugal), making them easier to detect consistently.

Lastly, a third set of mutations, *e.g.*, L5F, A27S, and N185D, were frequently found in clinical samples throughout the sampling period but only sporadically in wastewater samples. These mutations are also of low frequency but are almost exclusively associated with clinical samples. During this analysis, we also noticed that many of these mutations occurred in the BetaCoV_S1-NTD domain of the Spike protein.

We then assessed the density of mutations in each category (exclusive to clinical samples, exclusive to wastewater samples, or found in both sample types) within the functional and structural regions of the Spike protein (Figure 3D), taking into account also the horizontal coverage of each region (Figure 3E). We observed a coverage bias towards specific regions, particularly the superimposed Domain 2 – BetaCoV S1-CTD, and Region 7. Notably, these regions are those where we most frequently detect the same mutations in both sample types. Thus, two factors may influence our ability to detect mutations in wastewater samples through S gene sequencing: the coverage of the S gene and the mutation frequency.

### Wastewater sequencing reveals low-frequency mutations missed by clinical surveillance

We then directed our attention to missense SNPs exclusively found in wastewater samples. Interestingly, other studies have also identified mutations in wastewater that were not reported in clinical samples collected during the same period (Izquierdo-Lara et al., 2021, 2023; Smyth et al., 2022).

First, we investigated in worldwide clinical samples the presence of mutations identified exclusively in wastewater samples from Portugal and found that all, except two, have been previously identified, either in Portugal before our sampling period, or in other countries. Two mutations, G381R and Q607R, were detected in our wastewater samples 11.5 and 9.7 months, respectively, before being reported in Portuguese clinical samples. This highlights the power of WBS to detect low-frequency mutations, which is due to its intrinsic ability to sample a large population and capture the underlying viral genetic diversity. Additionally, the mutations D578E and T998K were never previously detected worldwide, emphasizing WBS’s ability to uncover novel mutations, which could potentially originate from unsampled infected individuals, viral tropism to different anatomical locations, such as the gastrointestinal tract, prolonged COVID-19 infection which potentiates in-host evolution, or even an animal reservoir (Gregory et al., 2022; Smyth et al., 2022).

We then investigated the distribution of wastewater missense SNPs not detected in our clinical samples across different countries (Figure 4A). It became apparent that some mutations emerged in multiple countries, while others were detected only in a few countries (Figure 4A). The countries with higher numbers of sequenced clinical samples (Figure 4A, bar chart in green) reported more mutations, and the mutations found at higher frequencies (Figure 4A, bar chart in blue) were identified across more countries. These results highlight the influence of mutation frequency on the ability to capture the full extent of viral genetic diversity and emphasize the importance of sequencing more samples to identify low-frequency mutations. Note that the mutations detected solely through wastewater S gene sequencing are present in very low frequencies in clinical samples worldwide (ranging from 2.09×10^-7^ to 8.55×10^-2^; Figure 4B), whereas they exhibit a diverse frequency range in our wastewater sample dataset (Figure 4C).

**Figure 4:**
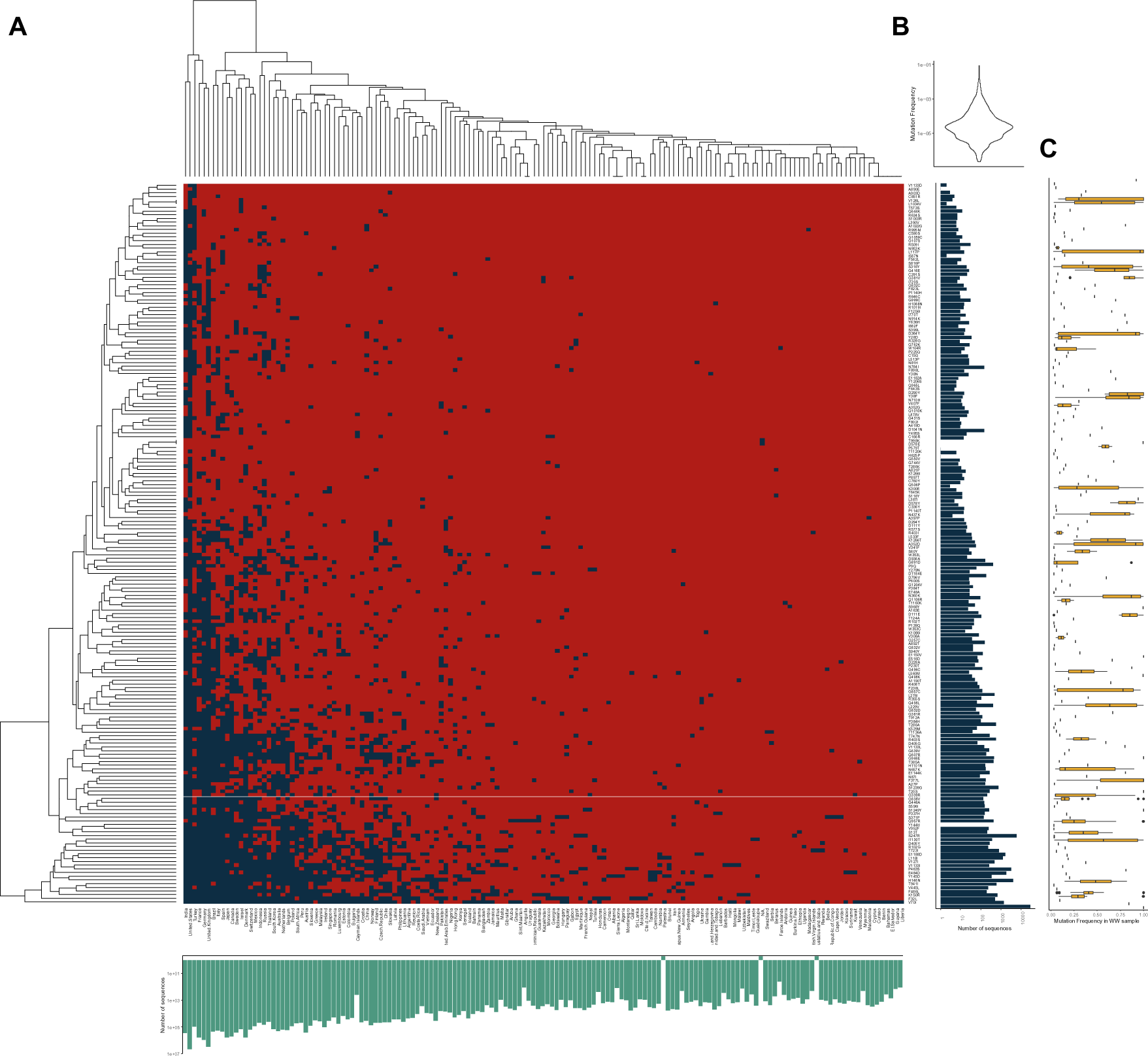
Amino acid substitution mutations in the S gene only identified in WW samples that were not identified in Portugal’s clinical samples. A. A heat map of occurrence (presence in blue, absence in red) of each mutation detected in WW samples from Portugal, absent from the clinical surveillance component in Portugal, but present in clinical samples from countries other than Portugal. Rows correspond to the mutations and columns to countries. Mutations and countries are sorted by the mean value of rows and columns, which is reflected in the clustering dendrograms. The green bar plot on the bottom of the heatmap shows the number of clinical SARS-CoV-2 sequences available for each country, highlighting the different sequencing efforts of each country. Palestine and Saint Eustatius and Saba are not considered as independent territories in GISAID, contrary to OutbreakInfo, and their sequencing efforts are included in Israel and The Netherlands, respectively. The blue bar plot of the right represents the number of sequences containing the mutation in the set of all available clinical sequences, highlighting that this set of mutations occurs rarely. B. Violin plot showing the distribution of the frequencies of these mutations in the represented countries, once again highlighting that these mutations occur in very low frequencies. C. Box plots representing the frequency of these mutations in the WWTPs analyzed in this study.

These findings highlight the need for an extensive sequencing effort to identify rare mutations, likely explaining why they have been overlooked in clinical surveillance in Portugal. It also underscores the importance of comprehensive clinical sampling to detect such infrequent mutations or alternatively, the adoption of WBS, which can efficiently track low-frequency mutations by sampling a larger population in one go.

### The S gene sequencing-based lineage assignment inadequately captures Omicron lineage progression

WBS gained traction during the COVID-19 pandemic, leading to the development of multiple tools for SARS-CoV-2 lineage assignment from challenging mixed sample sequencing data (Amman et al., 2022; Karthikeyan et al., 2022; Sapoval et al., 2023). The regular emergence of new lineages with increasing shared mutations challenges accurate lineage identification (Fontenele et al., 2021; Izquierdo-Lara et al., 2021; Swift et al., 2022). With over 300 Omicron sub-lineages, high-quality sequencing data becomes increasingly crucial for lineage assignment.

Given our sub-optimal sequencing data of the S gene from mixed samples containing multiple SARS-CoV-2 lineages, and the low horizontal coverage of a few samples, we required a robust bioinformatics tool for lineage assignment. Freyja pipeline demonstrated superior performance compared to other deconvolution methods (Baaijens et al., 2021; Katharina et al., 2022; Valieris et al., 2022) in terms of accuracy, false-positive rate, and computational efficiency, making it the selected method for this study.

We attempted lineage assignment in all 332 sequenced samples, and the results are summarized in Figure 1C. The resolution achieved by Freyja was not always sufficient to make assignments at the level obtained with the RT-PCR genotyping assays, specifically for the major Omicron lineages, leading to a high frequency of the ‘Others’ category. The limitations arise from the low and uneven S-gene horizontal coverage in our samples, with average values below 25% in several samples, along with the low number of samples analysed per week (Figure 1D), which collectively hinder the assignment success and the effective ability to capture the progression of sub-lineages and discriminate within.

Nonetheless, the major Omicron sub-lineages circulating in our sampling period (*i.e.*, BA.1.X, BA.2.X, BA.4.X, BA5.X, and XBB.X) have also been detected. BA.1.X was consistently present throughout the sampling period (Figure 1C), despite representing a residual frequency of SARS-CoV-2 infections according to clinical and genotyping assays (Figure 1A and 1B). BA.2.X was detected throughout the sampling period, gradually decreasing in frequency, particularly evident after the rise of XBB.X infections in week 5 of 2023. BA.4.X was always detected, but with higher proportions until week 46 of 2022 (Figure 1C), when it also decreased detection by both genotyping assays and clinical surveillance (Figure 1A and 1B). BA.5.X appeared throughout the sampling period (Figure 1C), aligning with clinical data as the dominant sub-lineage (Figure 1A). Its frequency gradually reduced with the rise of XBB.X sub-lineage. XBB.X was first identified by S gene sequencing in week 42 of 2022 (Figure 1C), coinciding with its first detection in clinical data (Figure 1A).

The results obtained from S gene sequencing-dependent lineage assignment agree with genotyping assays and clinical surveillance, but the progression of Omicron sub-lineages is not well captured by this methodology, likely due to coverage constraints in complex mixed-samples like wastewater.

## DISCUSSION

Genomic surveillance of SARS-CoV-2 played a crucial role in the COVID-19 pandemic, providing vital information for understanding virus transmission and evolution, and guiding effective control measures. However, with the success of vaccination campaigns leading to a decline in infections and their severity, the focus on genomic surveillance has gradually decreased worldwide since December 2022, and in Portugal since May 2022, when the country testing strategy changed. While understandable, given the current impact of the pandemic, this reduction leaves us unaware of the ongoing infection dynamics and emerging variants. As a result, more resource-efficient methods like WBS have been explored for population-wide genomic surveillance. To validate and optimize this new approach, it must be rigorously compared with current clinical surveillance methods, addressing challenges, and implementing necessary improvements.

Here, we conducted one year of wastewater-based surveillance on the Portuguese population, spanning from May 2022, when clinical surveillance sequencing efforts began to decline, until April 2023. The dynamics of SARS-CoV-2 were monitored using a combined approach of RT-PCR genotyping assays and targeted S gene sequencing.

Screening signature Omicron sub-lineage mutations via RT-PCR offers a fast and efficient method, providing community-level insights and adaptability to emerging mutations, making it a valuable addition to epidemiological surveillance. Moreover, it reduces costs compared to high-throughput sequencing, enabling broader implementation. However, this approach is limited to the identification of known sub-lineages with well-known diagnostic mutations and its discriminatory power diminishes with extensive Omicron VOC diversification. Additionally, targeted RT-PCR genotyping assays cannot detect new unknown lineages, low frequency mutations, or novel mutations. They also provide limited information about which variants are responsible for the overall increase or decrease of SARS-CoV-2 viral load in sewage due to their inability to determine lineage frequencies in these very complex samples. Thus, wastewater-based surveillance via RT-PCR genotyping alone cannot fully match the comprehensive information obtained from clinical genomic surveillance.

High-throughput sequencing methods offer significant advantages, providing a broader and less biased overview of viral genetic diversity and enabling detection of novel lineages, tracking low-frequency mutations, and identification of mutational hotspots. However, our data highlights the challenges inherent to wastewater samples, such as biological complexity, enzymatic inhibitors, viral RNA concentration and integrity, which are critical factors to obtaining high quality samples required for successful sequencing and full computational analyses (Izquierdo-Lara et al., 2021). Particularly evident is the impact introduced by samples in which S gene coverage is low, leading to an underestimation of mutational diversity and hindering lineage attribution, preventing the full reproduction of the Omicron sub-lineage succession patterns observed in clinical surveillance. As other authors already pointed out, in our work, no correlation was found between S gene coverage and RNA concentration or viral load (Izquierdo-Lara et al., 2021; Pérez-Cataluña et al., 2022), therefore other parameters such as wastewater storage, pre-process and concentration before nucleic acid extraction, or biases introduced by library construction and amplicon sequencing strategies, need to be analysed to infer their impact on RNA recovery and, consequently, on sequencing performance (Perez-Zabaleta et al., 2023; Tamáš et al., 2022). Genomics is a rapidly growing field that can yield useful information for public health surveillance, particularly during ongoing disease outbreaks. Many factors currently affect sequencing quality (*e.g.* coverage), influencing the full applicability of genomic data. Critical in the clinical and wastewater contexts is that not all available samples have the same potential to result in high quality sequencing outcomes, such as genome depth and coverage. Since horizontal coverage of the S gene may hinder the comprehensive detection of the wide spectrum of mutations circulating on a given population at the sewershed level, it is crucial to invest efforts alongside the entire genomics workflow, from sample collection and sample processing to library construction, sequencing technology and sequence data analyses, to obtain more accurate and representative outputs. Predictive models of resulting sequencing depth and coverage based on known variables associated with collected samples are thus in great demand.

The genotyping assays allowed the identification of mutation alleles found in one or more Omicron sub-lineages, frequently for longer periods compared to the clinical genomic surveillance data. This time lapse could be attributed to the larger population sampled in WBS, where one weekly wastewater sample represents up to 920000 people, whereas in clinical surveillance, an average of 134 persons were sampled per week. It can also reflect the less biased sampling approach in WBS, which includes both symptomatic and asymptomatic individuals, and the fact that in clinical surveillance, the presence of the virus is tested once (usually at the onset of infection), while in WBS it is detected for the full infection period. During that period, the ongoing excretion of viral RNA in faeces can persist approximately for an average duration of 14-21 days (Lescure et al., 2020; Pan et al., 2020), with SARS-CoV-2 shedding having been reported for up to 55 days in patients with complications or admitted to intensive care units (Lavania et al., 2022).

Another relevant aspect of our analysis concerns low-frequency mutations, including two that were first detected in wastewater samples and much later in clinical samples, and two mutations never previously reported worldwide. This is in line with other wastewater sequencing surveillance initiatives, where mutations in Spike were identified months before being reported in clinical samples (Izquierdo-Lara et al., 2023; La Rosa et al., 2023; Pérez-Cataluña et al., 2022), highlighting the potential of wastewater surveillance as an early warning system. For example, and considering the most recent variant of interest EG.5, a descendent of XBB.1.X (risk evaluation from 9 August 2023, by WHO), this lineage was identified in very low-frequency in our wastewater samples from week 16 of 2023, however it was reported on clinical data only from week 18 onward, re-enforcing the capability of WBS to anticipate trends and upsurges in a given region.

Early detection of low-frequency mutations is crucial to monitor the evolution of SARS-CoV-2, as they serve as the basis for natural selection and can give rise to new VOCs/lineages that may be more transmissible or cause more severe infections. We observed that several low-frequency mutations were detected in clinical samples but not in wastewater samples, likely due to their low frequency, making their detection more random in either sample type. On the contrary, we also found several mutations in wastewater samples that were not previously seen in Portugal through clinical surveillance. Similar findings were also made by Pérez-Cataluna and collaborators in Spain, and La Rosa and collaborators in Italy, with the report of exclusive wastewater mutations or mutations not reported as dominant in clinical samples (La Rosa et al., 2023; Pérez-Cataluña et al., 2022). The report of novel amino acid substitutions at Spike gene were also reported in other publications (Crits-Christoph et al., 2021; Pérez-Cataluña et al., 2021; Smyth et al., 2022).

The low-frequency and novel mutations may originate from unsampled individuals, which is not unlikely given that our sampling period corresponds to the time point in which clinical surveillance decreased in Portugal. They can also originate from asymptomatic cases, viruses replicating in different body sites (*e.g.*, respiratory *vs.* gastrointestinal tracts), prolonged infections leading to in-host evolution, or even from animal reservoirs whose excretions also end up in the same wastewater treatment plants (Gregory et al., 2022; Smyth et al., 2022).

Although not able to completely replace clinical surveillance, allelic discrimination assays and S gene sequencing can complement each other to overcome their respective limitations. We envision an implementation scheme in which the RT-PCR genotyping assays, selected based on the data being collected weekly and by the sequencing results, allow fast testing of a large number of samples, covering a very large set of the population. Additionally, successful implementation of S gene targeted sequencing in parallel will allow regular monitoring of virus evolution and detection, and tracking low-frequency mutations.

## CONCLUSION

In summary, the combination of wastewater genomic surveillance using RT-PCR assays and S gene sequencing provided useful insights into the diversity of circulating Omicron sub-lineages within the sampled population throughout 12 months. Although each method has its own limitations in terms of lineage discrimination, when used together, they offer a reliable snapshot of the local epidemiological situation. Additionally, this combined approach enables fast lineage attribution alongside the identification of low-frequency and novel mutations, which would typically require extensive clinical sampling and sequencing efforts. Further, they support the prolonged detection of lineages considered by clinical surveillance to be already out of circulation. The results generated in this work evidence the value of environmental surveillance to capture public health trends and underscore the effectiveness of WBS as a tracking system for virus variants. Moreover, WBS offers the opportunity to transform the cost per clinical sample into the cost per sewershed.

In conclusion, our findings emphasize that knowledge of the genetic diversity of SARS-CoV-2 at the population level can be extended far beyond via the combination of routine clinical genomic surveillance with wastewater sequencing and genotyping.

## Data Availability

All data produced in the present study are contained in the manuscript or are available upon reasonable request to the authors

## ACKNOWLEDGMENTS

We acknowledge the coordination efforts of AdP VALOR from the very first moment, particularly of Ana Katila Ribeiro, Marta Carvalho and Nuno Brôco, and the engagement of the Portuguese Environment Agency [APA] and the National Health Authority [DGS] as well, in setting up a national surveillance system. Thanks are due to Anabela Rebelo [APA].

We acknowledge the close collaboration of water utilities [AGERE, Águas do Algarve, Águas do Centro Litoral, Águas do Norte, Águas do Tejo Atlântico, Águas e Energia do Porto,

SIMDOURO, SMAS Almada] along this project and thank all their employees who contributed to wastewater sampling.

The institutional support of *Ministério do Ambiente e Ação Climática* and the European Commission’s *DG Joint Research Centre*, Directorate D – Sustainable Resources is gratefully acknowledged.

## DATA AND MATERIALS AVAILABILITY

Materials generated in this study will be made available upon request. All data needed to evaluate the conclusions of this work are present in the paper and/or the Supplementary Materials. This paper does not report original code. Any additional information required to reanalyse the data reported in this paper is available from the lead contact upon request.

## SUPPLEMENTARY MATERIALS

We provide supplementary files with supporting material, including Supplementary Table 1 and Supplementary Figures 1 to 5 (Figures S1-S5).

