## Supplemental Figures and Table for "Systematic SARS-CoV-2 S Gene Sequencing in Wastewater Samples Enables Early Lineage Detection and Uncovers Rare Mutations in Portugal"

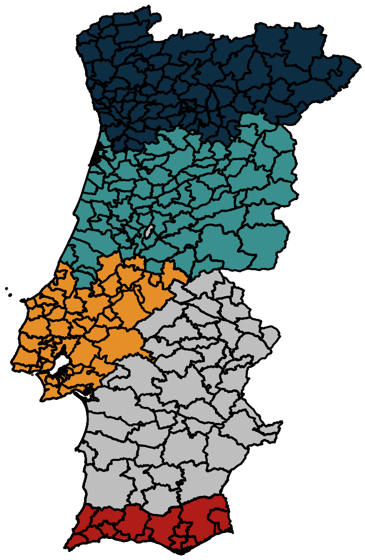

**Figure S1: The Portuguese Health Regions under analyses.** North, dark blue. Center, light blue. Lisbon and Tagus Valley, orange. Algarve, red.  Due to anonymization of data, the specific municipalities that were under monitoring in each region are not highlighted.

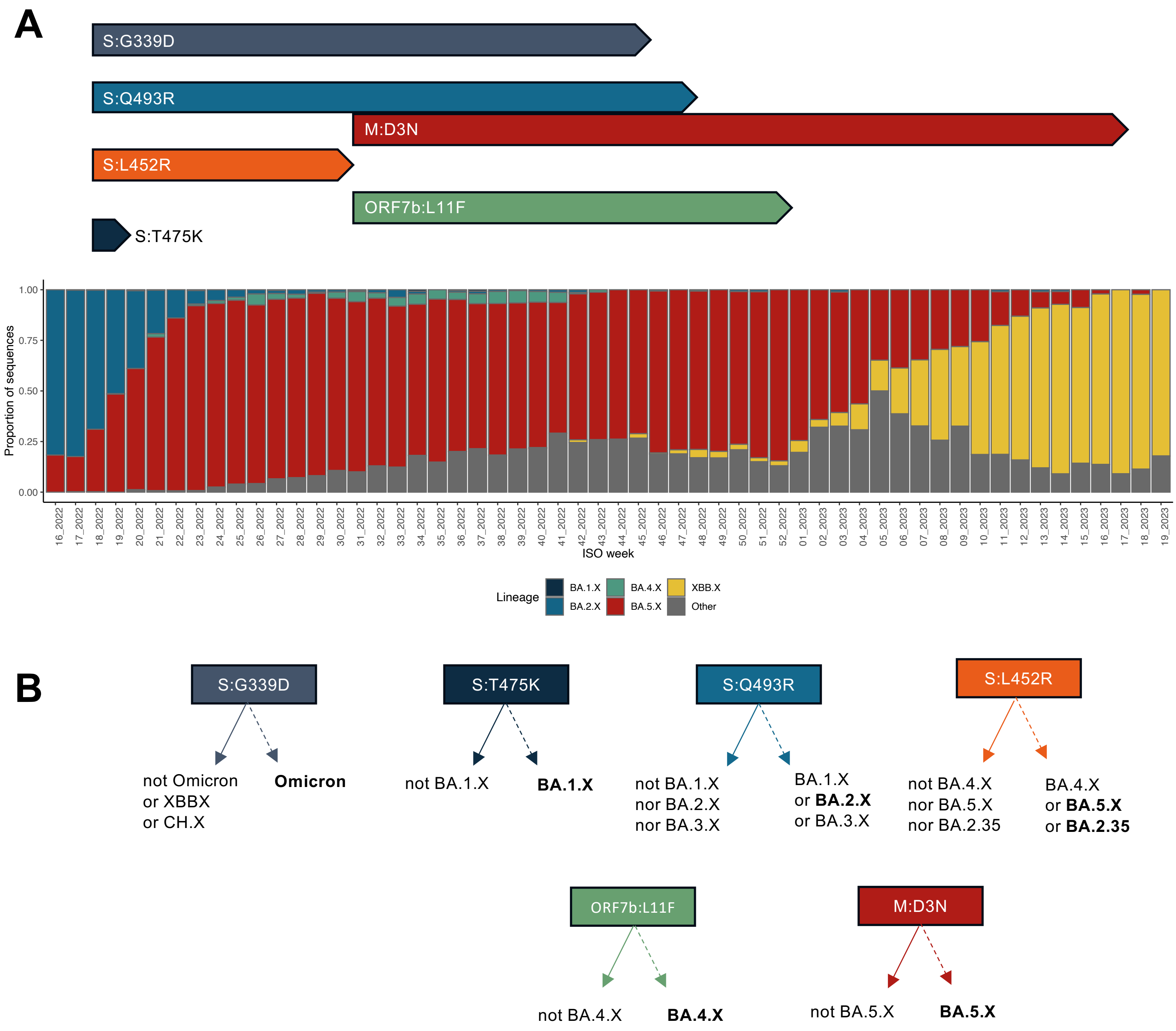


| Mutant allele | Omicron | BA.1.X | BA.2.X | BA.2.35 | BA.3 | BA.4.X | BA.5.X |
|---|---|---|---|---|---|---|---|
| S:G339D | X | | | | | | |
| S:T475K | | X | | | | | |
| S:Q493R | | X | X | X | X | | |
| S:L452R | | | | X | | X | X |
| ORF7b:L11F | | | | | | X | |
| M:D3N | | | | | | | X |

**Figure S2: Combination of RT-PCR Genotyping Assays for Identifying Specific Omicron Sub-lineages. A.** The bar graph illustrates the proportion of clinically derived sequences from Portugal obtained from GISAID, categorized by the major sub-lineages listed in the color-coded legend. Aligned with the weeks on the horizontal axis of the bar chart, the time span during which the genotyping assays were applied is represented, each identified by the specific mutation they detect. **B.** Graphical and tabular representation of the lineage assignment algorithm based on the RT-PCR data. Dashed lines represent the detection of the mutant allele, while filled lines represent the detection of the wild-type allele. On the table, the X marks the amplification of the mutant allele in each assay.

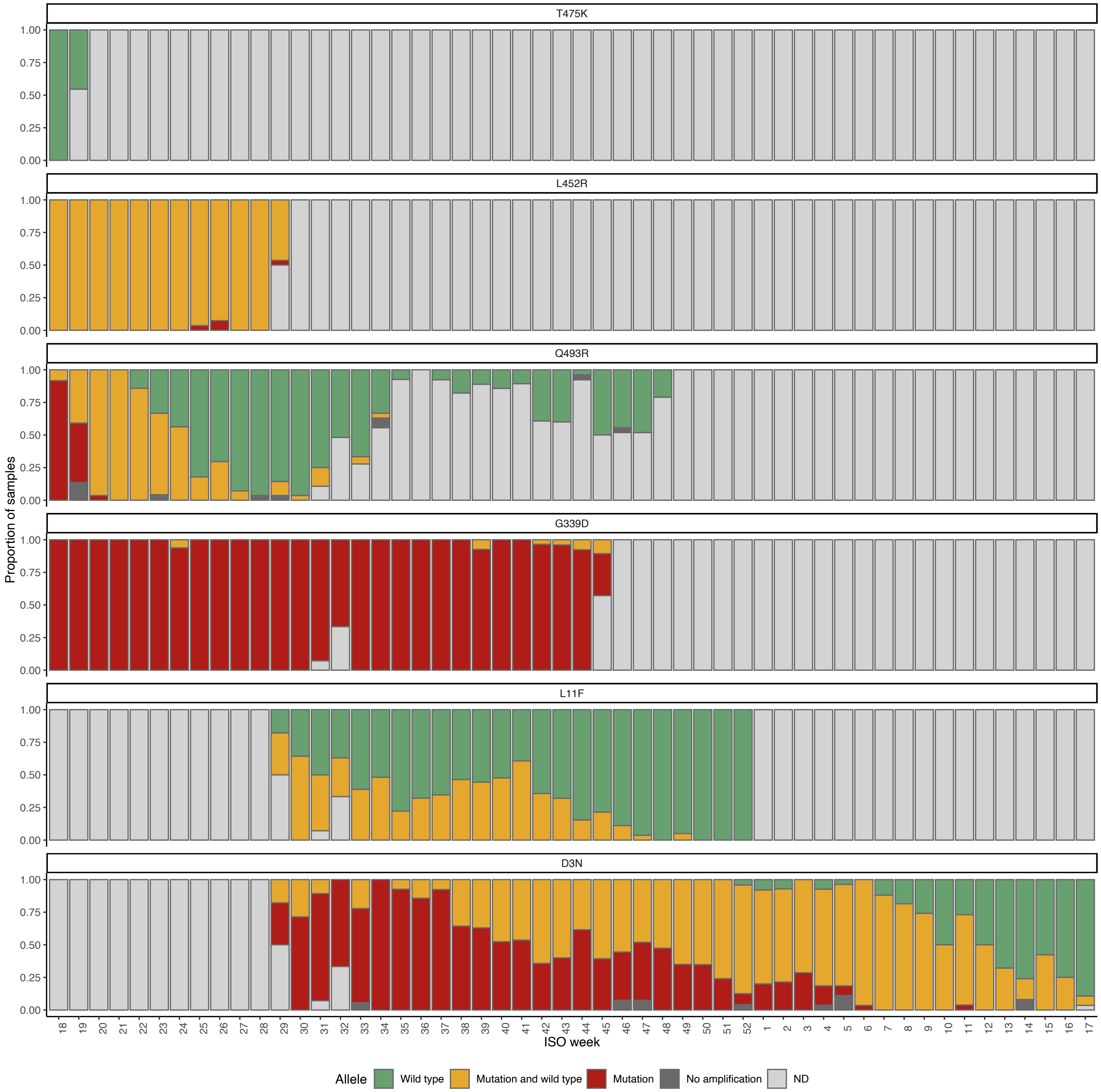


**Figure S3: Results of RT-PCR Allelic Discrimination Assays for Identifying Major Omicron Sub-lineages.** The bar plot displays the proportion of samples for each assay (identified by the mutation they detect) in which amplification occurred solely for the wild-type allele (green), only the mutated allele (red), or both (orange). Dark grey represents samples with no amplification, while light grey indicates samples not tested in a given week. Weeks are identified by ISO code, with those between 18 and 52 corresponding to the year 2022, and those from 1 to 17 representing 2023. ND – Not Determined.

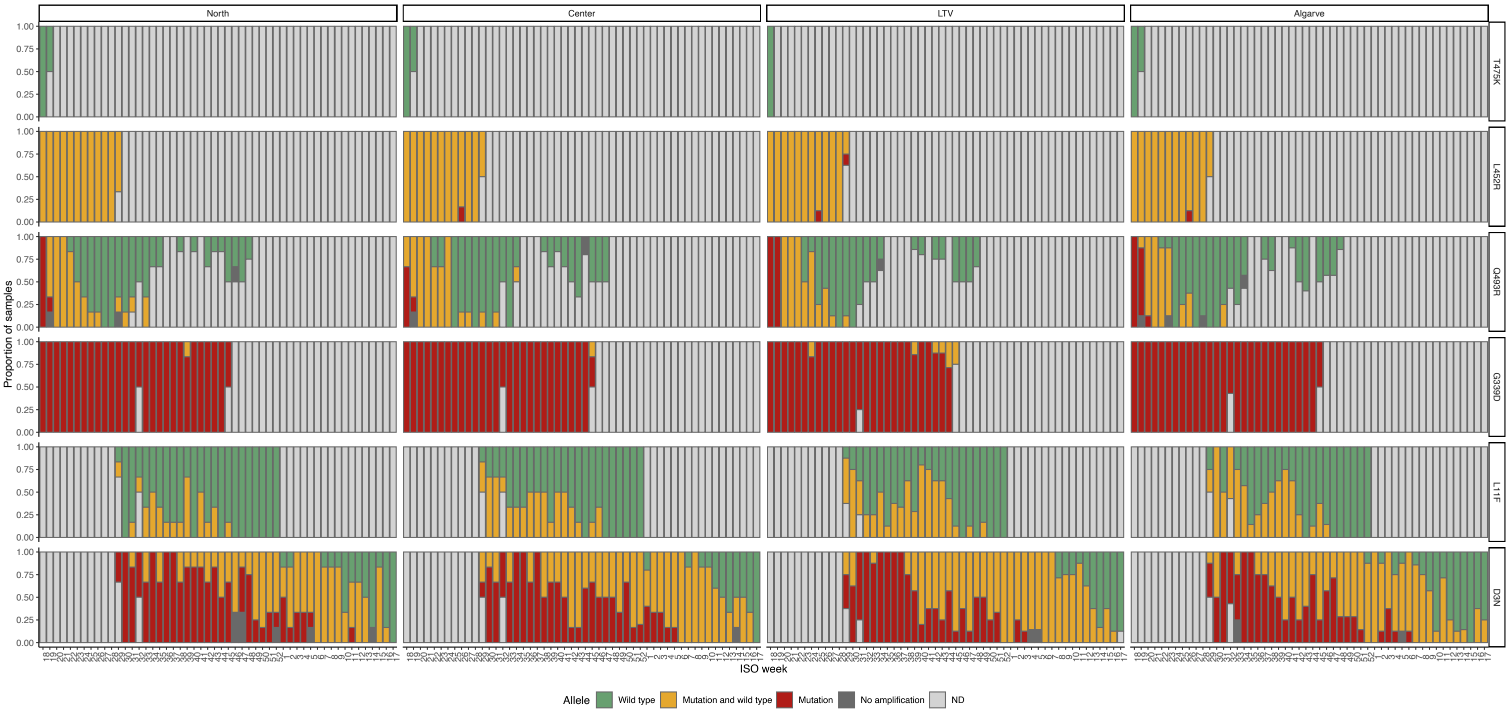


**Figure S4: Results of RT-PCR Allelic Discrimination Assays for Identifying Major Omicron Sub-lineages by Health Regions.** In this chart are represented the same data as in Figure S2 but stratified by region. The bar graph displays the proportion of samples for each assay (identified by the mutation they detect) in which amplification occurred solely for the wild-type allele (green), only the mutated allele (red), or both (orange). Dark grey represents samples with no amplification, while light grey indicates samples not tested in a given week. Weeks are identified by ISO code, with those between 18 and 52 corresponding to the year 2022, and those from 1 to 17 representing 2023. LTV, Lisbon and Tagus Valley. ND - Not Determined.

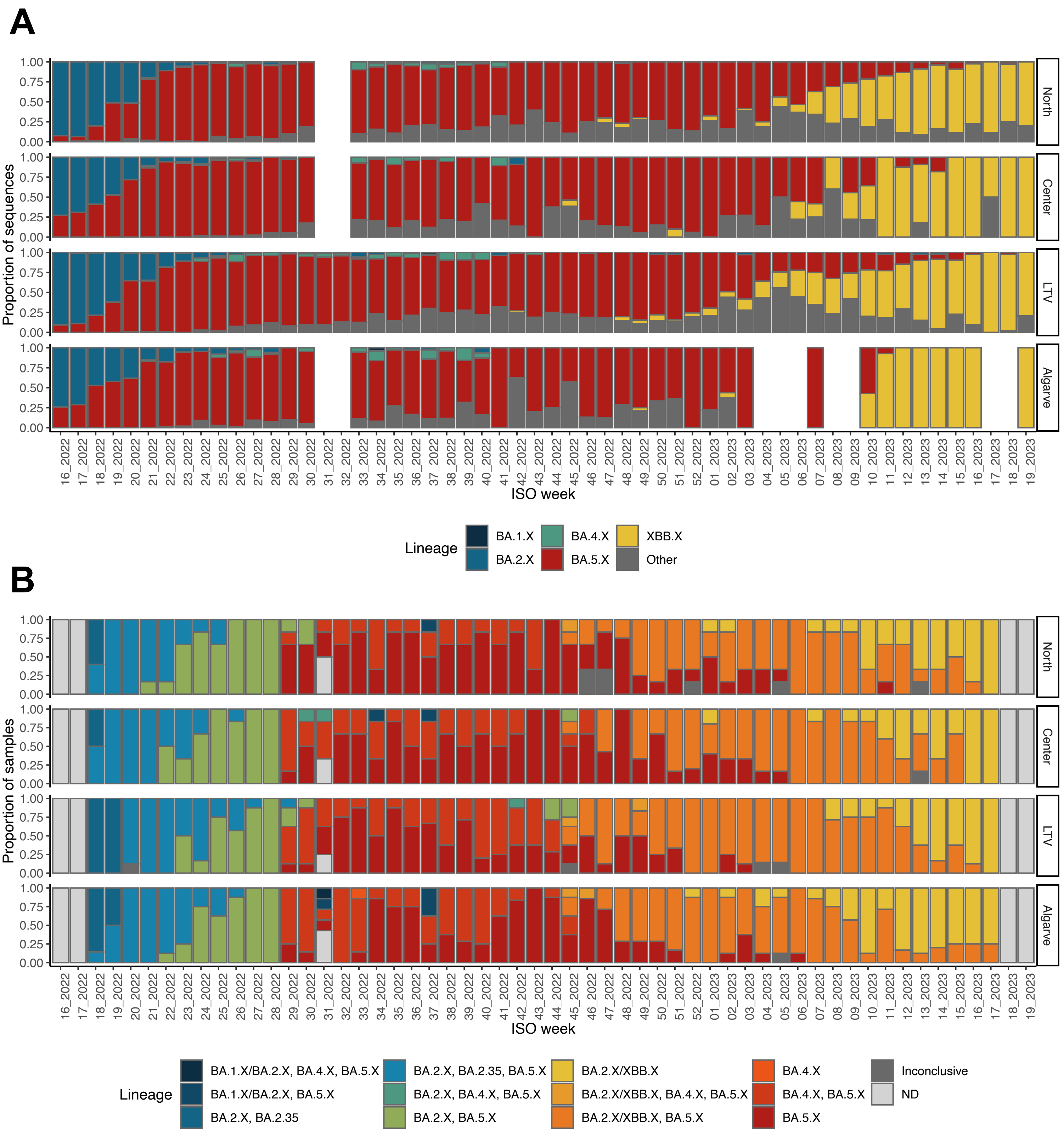


**Figure S5: Lineage Frequency Evolution by Health Regions. A.** Lineage assignment of sequences derived from clinical samples collected in Portugal, within North, Center, Lisbon and Tagus Valley, and Algarve Health Regions, from ISO week 16 of year 2022 to ISO week 19 of year 2023. Descendant lineages of the five major circulating lineages (BA.1, BA.2, BA.4, BA.5, and XBB) were combined into a single category for simplicity. The remaining (minor) lineages were grouped into the Other category. **B.** Lineage assignments in WWTP samples based on real-time PCR genotyping assays. The samples were grouped into 14 categories based on the discrimination power of the genotyping assays. Most of the categories grouped samples with multiple major circulating lineages detected. The forward slash indicates that the test cannot distinguish between the two lineages. In a few samples, amplification was not obtained in all necessary real-time PCR assays, leading to grouping them in the category "Inconclusive," or they were not tested due to lack of quality RNA, and these samples were grouped in the ND category. LTV represents Lisbon and Tagus Valley region. ND – Not Determined.

**Supplementary Table 1.** Specificities of the allele specific RT-PCR genotyping assays

| Genotyping Assay | Mutation[1] | ThermoFisher Scientific assay ID |
|---|---|---|
| **G339D** | S.G339D.GGT.GAT | CV47VRX |
| **L452R** | S.L452R.CTG.CGG | A51819 |
| **Q493R** | S.Q493R.CAA.CGA | CVH49P2 |
| **T547K** | S.T547K.ACA.AAA | CVYMJGA |
| **D3N** | M.D3N.GAT.AAT | CVAAAAK |
| **L11F** | ORF7b.L11F.TTG.TTT | CVCE3VH |

[1]Mutation name convention: Gene.Mutation.Reference Codon.Mutant Codon
